# Mapping the emergence of SARS-CoV-2 Omicron variants on a university campus

**DOI:** 10.1101/2022.04.27.22274375

**Authors:** Ana A. Weil, Kyle G. Luiten, Amanda M. Casto, Julia C. Bennett, Jessica O’Hanlon, Peter D. Han, Luis Gamboa, Evan McDermot, Melissa Truong, Geoffrey S. Gottlieb, Zack Acker, Caitlin R. Wolf, Ariana Magedson, Eric J. Chow, Natalie K. Lo, Lincoln C. Pothan, Devon McDonald, Tessa Wright, Kathryn McCaffrey, Marlin D. Figgins, Janet A. Englund, Michael Boeckh, Christina M. Lockwood, Deborah A. Nickerson, Jay Shendure, Trevor Bedford, James P. Hughes, Lea M. Starita, Helen Y. Chu

**Affiliations:** Department of Medicine, University of Washington, Seattle, WA; Vaccine and Infectious Diseases Division, Fred Hutchinson Cancer Research Center, Seattle, WA; Brotman Baty Institute, Seattle, WA; Department of Genome Sciences, University of Washington, Seattle, WA; Environmental Health & Safety Department, University of Washington, Seattle, WA; Department of Global Health, University of Washington, Seattle, WA; Seattle Children’s Research Institute, Department of Pediatrics, University of Washington, Seattle, WA; Department of Laboratory Medicine and Pathology, University of Washington, Seattle, WA; Howard Hughes Medical Institute, Seattle, WA; Department of Biostatistics, University of Washington, Seattle, WA

## Abstract

Novel variants continue to emerge in the SARS-CoV-2 pandemic. University testing programs may provide timely epidemiologic and genomic surveillance data to inform public health responses. We conducted testing from September 2021 to February 2022 in a university population under vaccination and indoor mask mandates. A total of 3,048 of 24,393 individuals tested positive for SARS-CoV-2 by RT-PCR; whole genome sequencing identified 209 Delta and 1,730 Omicron genomes of the 1,939 total sequenced. Compared to Delta, Omicron had a shorter median serial interval between genetically identical, symptomatic infections within households (2 versus 6 days, P=0.021). Omicron also demonstrated a greater peak reproductive number (2.4 versus 1.8) and a 1.07 (95% confidence interval: 0.58, 1.57; P<0.0001) higher mean cycle threshold value. Despite near universal vaccination and stringent mitigation measures, Omicron rapidly displaced the Delta variant to become the predominant viral strain and led to a surge in cases in a university population.

## Introduction

Persistent SARS-CoV-2 circulation has led to the continued emergence of variants of concern (VOCs). On November 26, 2021, the World Health Organization designated Pango lineage B.1.1.529 as Omicron, a VOC which rapidly spread globally. Omicron is classified into sublineages BA.1, BA.2, and BA.3, etc., while BA.1 and BA.2 have several designated sublineages^1^. Mutations of the Omicron variant have demonstrated enhanced transmissibility despite widespread population immunity, as evidenced by the exponential increase in cases over shorter time periods compared to prior VOCs^2–4^. There is also population-level, genomic, and *in vitro* evidence of decreased vaccine effectiveness against Omicron compared to the Delta variant and of partial evasion of vaccine-induced immunity by Omicron, leading to high numbers of breakthrough infections^5–8^. Studies have shown mixed results on differences in Omicron viral load compared to the Delta variant, with evidence of either lower or comparable viral loads for Omicron^9–12^. Omicron household transmission has been reported to have a higher attack rate and lower serial interval compared to Delta, although the majority of studies to date have not used genomic data to assess the serial intervals in intra-household transmission^13–19^. There remain gaps in our understanding of the transmission dynamics and molecular epidemiology of VOC emergence in US populations.

Throughout the COVID-19 pandemic, university campuses have been sites of SARS-CoV-2 outbreaks^20–23^. Many universities provide free, convenient testing to facilitate SARS-CoV-2 surveillance within campus communities^20,22,24^. Using data collected from September 2021 to February 2022 through a campus testing program, we describe the rapid emergence of Omicron in a highly vaccinated university community, and the clinical characteristics and transmission dynamics of the Omicron variant compared to the Delta variant. We used molecular epidemiology to track emergence of variants and examine intra-residence infections in congregate living settings.

## Online Methods

The Husky Coronavirus Testing (HCT) research study provides SARS-CoV-2 testing at the University of Washington (UW), a large public university in Seattle, Washington, USA^20^. University-wide mitigation policies during this study period included indoor masking, air filtration, limitations in size of gatherings, and mandatory vaccination for faculty, staff, and students, resulting in completion of the primary vaccine series for 98.6% of students, 98.9% of staff, and 99.7% of faculty by January 2022^25^. Individuals were eligible to enroll in the study if they were faculty, staff, or students at the university and were English-speaking. Clinical symptoms and vaccination status were collected through electronic questionnaires. Participants completed a daily attestation via email or text message, and those who reported new symptoms, exposure to a known SARS-CoV-2 case, or recent travel were offered SARS-CoV-2 testing. Additionally, participants could request testing for any reason. Data were collected using Project REDCap^26,27^.

### Swab collection

Testing was performed through three mechanisms: observed self-swab at a staffed kiosk, unobserved self-swab returned to a campus testing dropbox, or unobserved self-swab returned to the laboratory via courier^28^. Two swab types were used; a US Cotton #3 swab (SteriPack Polyester Spun Swab), returned in a 10mL tube, was used for all unobserved collection returned via courier, and for some observed collection testing at times of supply chain issues. The RHINOstic^TM^ Automated Nasal Swab (Rhinostics RH-S000001), returned in a Matrix^TM^ 1.0mL ScrewTop Tube (Thermo Fisher 3741), was used for observed kiosk and unobserved dropbox swab collections.

### Laboratory methods

All swabs were stored dry, with no preservative or media, and eluted with 1mL Tris-EDTA for US Cotton #3, or 300µL Tris-EDTA for RHINOstic^TM^. 50µL of eluate was treated with proteinase K and heat for direct RT-qPCR (Swab-Express RT-qPCR) as previously described^29^. The RT-qPCR assay employs custom probe sets for SARS-CoV-2 Orf1b and S gene designed against the ancestral strain that are multiplexed with a probe set for human RNase P^29^. Briefly, 5uL of prepared eluate was transferred to four multiplexed RT-qPCR reactions, two Orf1b-FAM plus RNase P-VIC and two S-FAM plus RNase P-VIC. Positive samples had SARS-CoV-2 targets detected in three or four reactions and an internal control RNase P amplification detected in at least three reactions; however, the 157-158 deletion in Delta variants results in S gene target failure or delay in our assay.

### Genomic sequencing

Viral genome sequencing was attempted on SARS-CoV-2 positive specimens with a high quantity of SARS-CoV-2 RNA, generally having Orf1b cycle threshold (Ct) ≤30. Nucleic acids were extracted (Magna Pure 96, Roche) and sequencing libraries prepared (Illumina COVIDSeq kit). Genomes were sequenced (Illumina NextSeq2000 P200 kit) and consensus genomes were assembled against the SARS-CoV-2 reference genome Wuhan/Hu-1/2019 (Genbank accession MN908947) using a modified iVar pipeline^30^. Consensus sequences were deposited to GenBank and GISAID (see Supplementary Materials). We considered “BA.1” to include the parental lineage and all BA.1 sublineages, and “BA.2” to include the parental lineage and all BA.2 sublineages.

### Statistical Analysis

We used the term “infection date” to describe the collection date of each person’s first SARS-CoV-2 positive sample, and to represent the first known date of infection regardless of symptom status. For participants who tested positive for SARS-CoV-2 more than once between September 10, 2021 and February 14, 2022, the first infection was included in our analysis. The proportion of cases reporting various symptoms were compared by variant using Pearson’s chi-squared tests. Median serial interval of symptomatic participants in clusters were compared by variant using a non-parametric Mann-Whitney U test.

COVID-19 vaccination status was collected at enrollment, updated monthly, and updated at or after collection of the SARS-CoV-2 positive samples. Participants self-reported vaccine manufacturer name, dose number, and date of receipt. Vaccination status for participants is dynamic, and in this manuscript the term “vaccination status” reflects the status on the date a positive swab was taken. Fully vaccinated was defined as completion of the primary series at least two weeks prior to the positive test date. Partially vaccinated was defined as an incomplete two-dose primary series or less than two weeks since completion of the primary series. Unvaccinated was defined as confirmed no vaccination. Vaccination was defined as unknown for participants who reported invalid dates or no information at all. A participant was considered boosted if they received a booster dose at least two weeks prior to the positive test date, partially boosted if fewer than two weeks, and not boosted if no booster was received by the positive test date.

Shared residence was defined as the same apartment, dorm room, or unit number, or by the same street address for single-unit residences. Clusters of positive cases were defined as living within a shared residence with identical SARS-CoV-2 sequences. An index date and serial interval were calculated for each cluster with at least two symptomatic individuals. Serial interval was defined for each non-index individual in a cluster as the duration of time between the index symptom onset date to non-index individual’s symptom onset date. Symptom onset date was defined as the earliest symptom onset date within one week before or after each individual’s positive test. Individuals were considered asymptomatic if they reported no symptoms within one week before and after testing positive.

Multiple linear regression models were used to estimate mean difference in Ct between Omicron and Delta variant cases, adjusting for age, symptom status (symptomatic versus asymptomatic), average RNase P gene value, days since symptom onset among those with symptoms, and vaccination status (primary series vs. booster) and days since last COVID-19 vaccination among those fully vaccinated. Additionally, we estimated the mean difference in Ct between Omicron lineages BA.1 and BA.2. Regression analyses were restricted to RHINOstic^TM^ swabs, due to previously observed differences in Ct between RHINOstic^TM^ swabs and US Cotton #3 swabs^20^. Mean Ct was calculated using only Orf1b Ct values due to differences in S-gene amplification between Delta and Omicron. Analyses were conducted in R (R-4.1.1, R Core Team, 2021).

Maximum likelihood phylogenetic trees were constructed using the Nextstrain Augur software package^31^ using default parameters for SARS-CoV-2 as outlined on the Nextstrain GitHub webpage^32^. Nextstrain Auspice software was used for tree visualization. Our genomic analyses also included publicly available SARS-CoV-2 genomes for other Washington state samples from the GISAID EpiCoV database^33^, which were screened using Nextclade version 1.10.0^34^. Any sequences deemed to be of “bad” or “mediocre” quality by this tool (due to missing data, mixed sites, private mutations, mutation clusters, frameshifts, or stop codons) were excluded from further analyses.

To estimate the number of Delta viral introduction events onto campus represented by the 209 sequenced Delta samples, we created a tree that included these samples along with all GISAID Delta genomes from samples collected in Washington state between September 1, 2021, and February 14, 2022, meeting our quality criteria (N=15,406). The Nextstrain Augur “traits” subcommand was used to infer campus versus non-campus states for all internal nodes. An introduction event was presumed to have occurred in all cases in which a Washington state non-university campus parent node connected with an on-campus child node. To assess the accuracy of this estimate given the number of Washington state Delta genomes available, we re-calculated the introduction event number using 15 subsamples of the total pool of available non-study genomes varying in size from N=1,000 to N=15,000. We repeated this process with a tree which included the 1,730 sequenced Omicron samples and all GISAID Omicron samples that were collected in Washington state before February 14, 2022 and met quality criteria (N=14,359) to obtain an estimate of the number of Omicron introduction events onto campus; the accuracy assessment for this estimate was done using 14 subsamples of Washington state Omicron genomes varying in size from N=1,000 to N=14,000.

### Transmission dynamics

Using all sequenced study samples and overall daily case counts over the study period, we estimated variant-specific effective reproduction numbers (R_t_) for Delta and Omicron. To do this we reconstructed variant-specific incidence from observed daily variant proportions using a multinomial likelihood for variant proportion and negative binomial likelihood for cases. This reconstructed incidence was used to compute the effective reproduction number for Delta and Omicron while reflecting the observed shorter serial interval of Omicron versus Delta^35^.

The UW IRB approved this study (#00011148).

## Results

A total of 37,985 participants were enrolled as of February 14, 2022. 74,995 samples were collected from 24,393 participants between September 10, 2021, and February 14, 2022. A total of 3,630 samples (4.8%) were SARS-CoV-2 positive, representing 3,048 individuals. Genomic sequencing of 2,101 samples from 1,939 individuals identified 209 Delta and 1,730 Omicron cases (**Figure 1**). Six individuals had both sequenced Delta and Omicron infections during the study period; only the first infection was considered for each individual.

**Figure 1:**
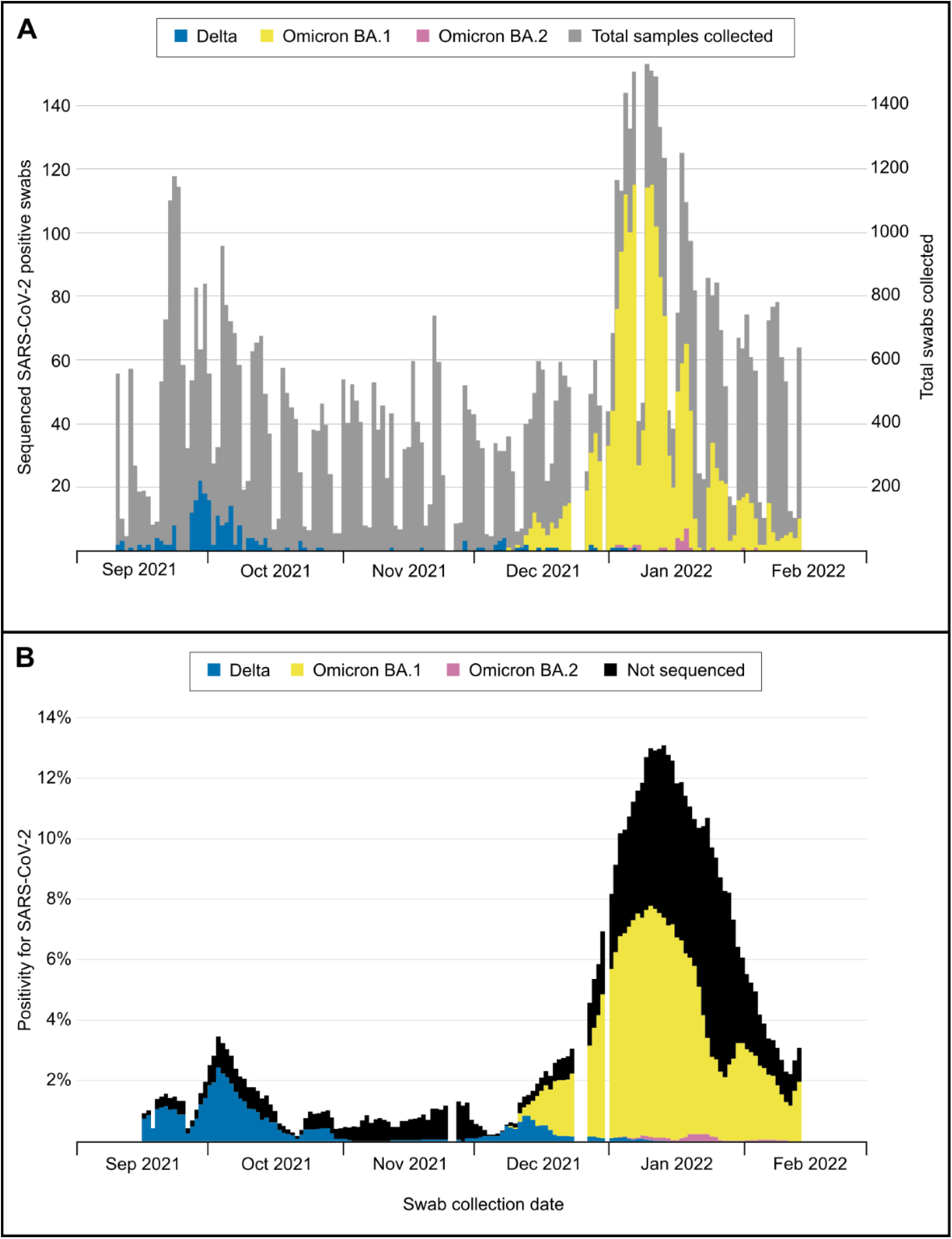
Sequenced SARS-CoV-2 positive samples collected from September 10, 2021 through February 14, 2022, by Pango Lineage. (A) Daily counts of total samples collected and positive samples. (B) SARS-CoV-2 7-day average percent positivity. Testing demand was reduced on weekends, and operations were paused for holidays (represented by gaps in testing), inclement weather, and campus closures.

### Clinical characteristics

The median age of participants with infection was 20 years (range 18-66) for Delta and 21 years (range 17-79) for Omicron (**Table 1**). Most SARS-CoV-2 cases were among students (90.9% of Delta cases, compared to 87.9% of Omicron cases). Residing in a household with a density of ≥6 was reported for 34.0% of Delta and 23.8% of Omicron cases. 18.2% of Delta and 18.3% of Omicron cases were asymptomatic at the time of swabbing. Among symptomatic cases, the most reported symptoms were rhinorrhea/congestion (69.6% and 62.4% for Delta and Omicron, respectively), cough (59.1% and 61.5%), and sore throat (56.1% and 69.2%). Loss of sense of taste or smell was more common among Delta cases (11.1% of those with Delta vs. 2.8% of those with Omicron, P<0.001). Myalgias, fever, and chills were more prominent in Omicron cases (29.8%, 34.1%, and 24.4%, respectively) than Delta (15.8%, 25.1%, and 15.8%; P<0.001, P=0.025, and P=0.015). The mean time from symptom onset to first positive sample was 2.82 days (standard deviation (SD): 2.03) for Delta and 2.76 days (1.93) for Omicron.

**Table 1.**
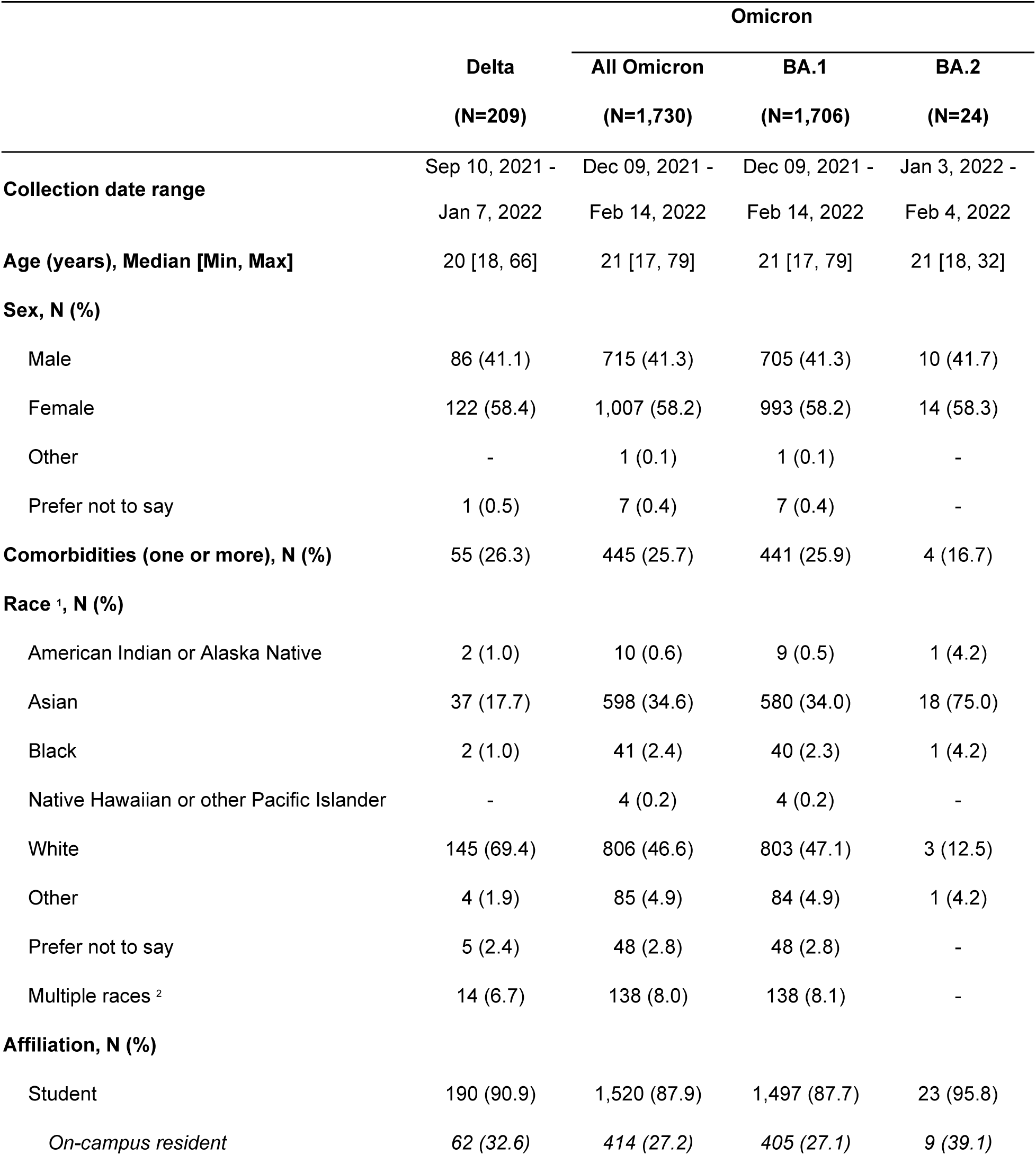
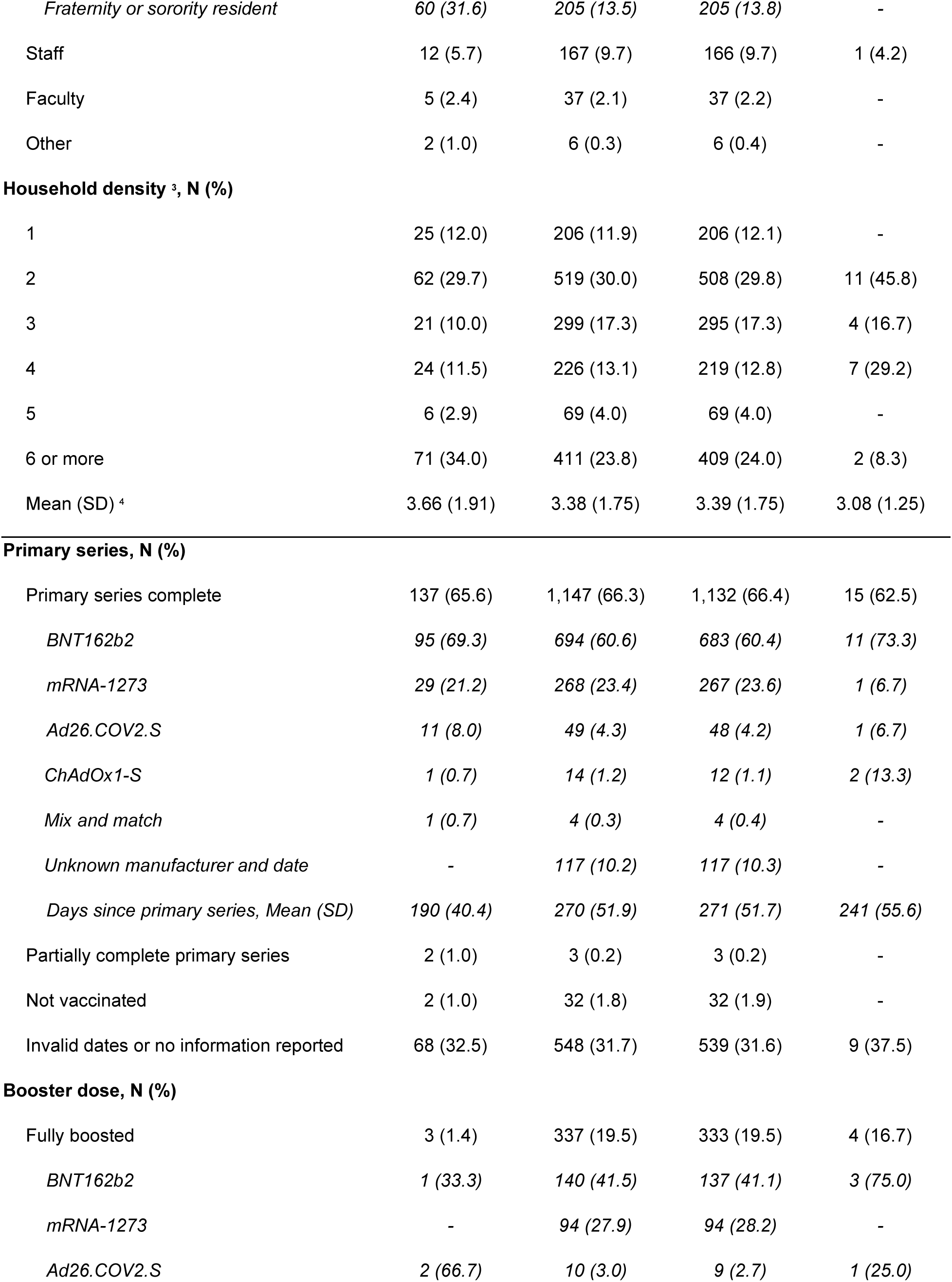
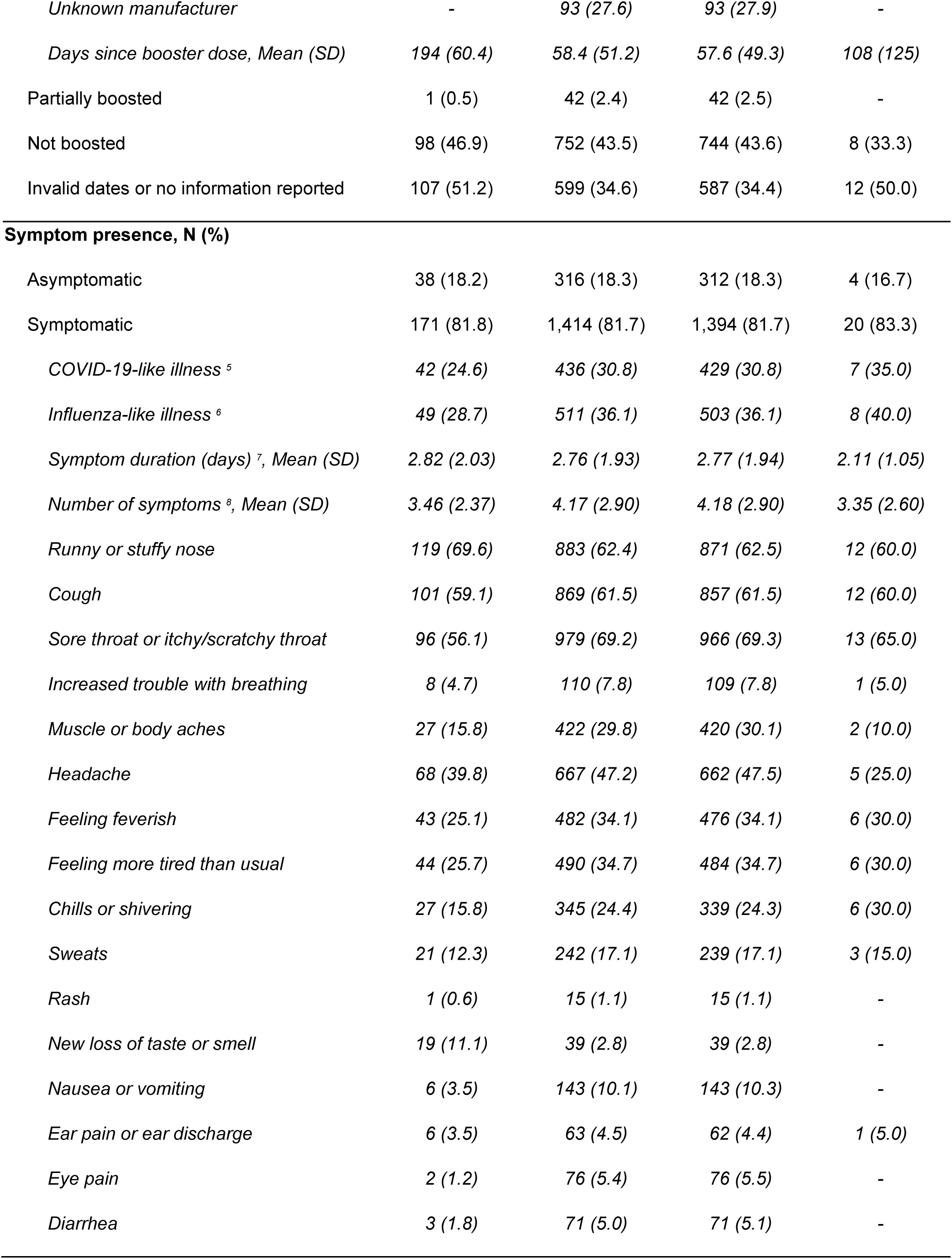
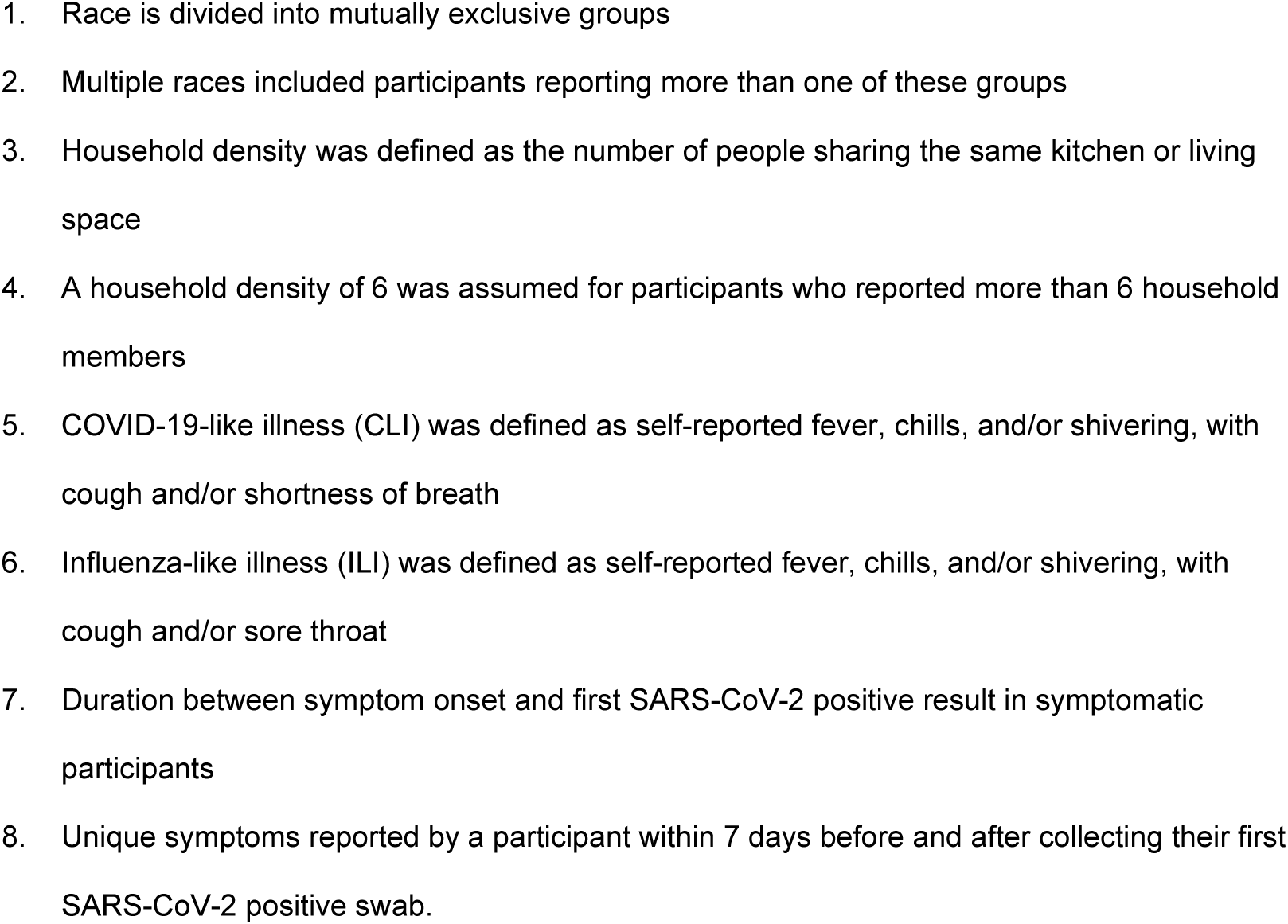
Demographic characteristics and symptom profiles for Delta and Omicron variant infected study participants, September 10, 2021, through February 14, 2022.

COVID-19 vaccination status was known at the time of infection for 141 (67.5%) of Delta and 1,182 (68.3%) of Omicron cases. For those with known vaccination status, 1,147 (97.0%) with Delta and 137 (97.1%) with Omicron completed a primary series with an additional 2 (1.4%) with Delta and 3 (0.3%) with Omicron that partially completed the primary series. Three (2.1%) with Delta and 337 (28.5%) with Omicron received a booster dose at least two weeks before infection and 1 (0.7%) with Delta and 42 (3.6%) with Omicron received a booster dose less than two weeks before infection. Two (1.4%) with Delta and 32 (2.7%) with Omicron were unvaccinated. Intervals between infection and last mRNA vaccine dose received are shown by vaccination status and variant in **Figure 2**. Most vaccinated participants completed their primary series by early Spring 2021 and days since primary series for Omicron cases (median 271 days, IQR: 251, 292) were higher than for Delta cases (median 194 days, IQR: 169, 224).

**Figure 2.**
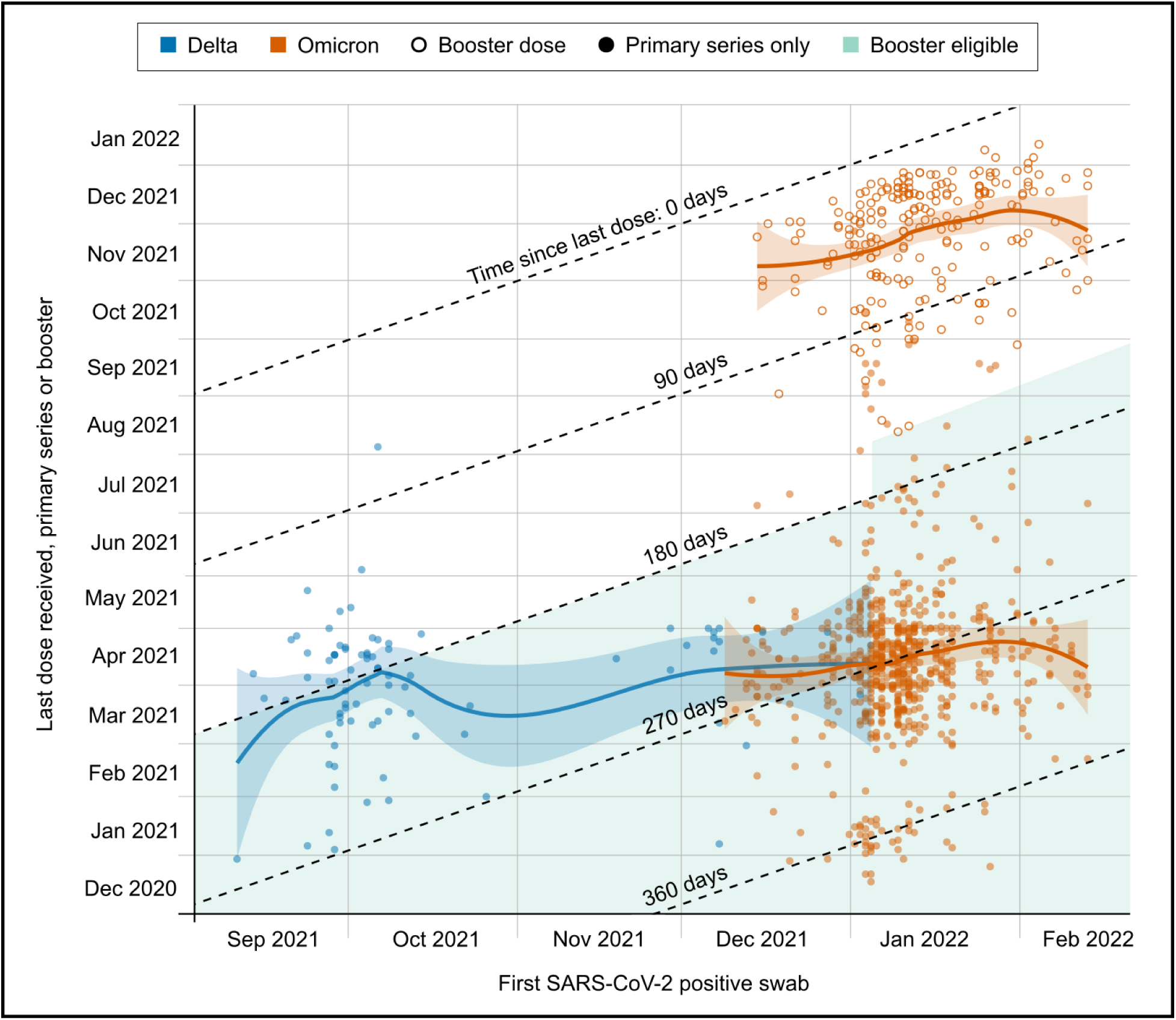
Interval between Delta or Omicron infection and last mRNA vaccine dose received. Loess curves with shaded 95% CIs shown for each variant and by booster status for Omicron. Period of booster eligibility is 180 or 150 days (beginning January 4, 2022) after primary series completion. In the US, a booster dose of BNT162b2 was available with limited eligibility on Sept 25, 2021, mRNA-1273 and Ad26.COV.2.S on Oct 20, 2021, and general eligibility on Nov 21, 2021. Not shown are participants who were unvaccinated, partially vaccinated, had unknown vaccination status, or received a vaccine other than BNT162b2 or mRNA-1273.

In our Ct value analysis, we compared the first positive, sequenced sample from each individual detected using our standard swab type (RHINOstic^TM^ swabs) (N=1,870, excluding 27 Delta and 42 Omicron cases detected using US Cotton #3 swabs). Adjusting for age, symptom status, and average RNase P gene value, the mean Orf1b Ct was 1.07 higher (95% confidence interval 0.58, 1.57; P<0.00001) among Omicron compared to Delta cases. Mean adjusted difference in Orf1b Ct comparing symptomatic to asymptomatic cases was –1.11 (95% CI, -1.50, –0.74; P<0.00001) and for each 1-unit increase in average RNase P gene value was 0.39 (95% CI, 0.35, 0.43; P<0.00001). Results did not change in a sensitivity analysis without adjustment for symptom status (data not shown). Among symptomatic individuals (N=1,466), days since symptom onset was significantly associated with a higher Ct value (0.29 higher per day, [95% CI: 0.20, 0.38], P<0.00001) and therefore lower semiquantative viral loads were observed in those with a longer duration of symptoms at the time of sample collection (**Table 2**). We did not find a difference in semiquantative viral load comparing Omicron Pango lineages BA.1 and BA.2 (N = 1,688, **Supplemental Table 1**).

**Table 2.** Cycle threshold comparisons by Delta and Omicron variants.

|  | Mean unadjusted<br>difference in Orf1b Ct<br>value (95% CI) | p-value | Mean adjusted<br>difference in Orf1b Ct<br>value (95% CI)* | p-value |
| --- | --- | --- | --- | --- |
| **All Delta and Omicron positive individuals, adjusted for age, symptoms, and average RNase P gene value**<br>**N=1,870 (Delta = 182, Omicron = 1688)** |  |  |  |  |
| Variant (Omicron vs. Delta) | 1.30 (0.76, 1.84) | **<0.00001** | 1.07 (0.58, 1.57) | **<0.00001** |
| Age (years) | -0.01 (-0.03, 0.01) | 0.29 | -0.01 (-0.03, 0.01) | 0.22 |
| Symptoms (symptomatic vs. asymptomatic) | -0.94 (-1.35, -0.53) | **<0.00001** | -1.11 (-1.50, -0.74) | **<0.00001** |
| Average RNase P gene value | 0.39 (0.35, 0.43) | **<0.00001** | 0.39 (0.35, 0.43) | **<0.00001** |
| **Symptomatic individuals (with symptom onset on or before day of swab) adjusted for age, days since symptom onset, and average RNase P gene value**<br>**N=1,466 (Delta = 144, Omicron = 1322)** |  |  |  |  |
| Variant (Omicron vs. Delta) | 1.08 (0.48, 1.68) | **0.0004** | 0.81 (0.26, 1.37) | **0.004** |
| Age (years) | -0.01 (-0.03, 0.01) | 0.44 | -0.01 (-0.03, 0.01) | 0.29 |
| Days since symptom onset | 0.32 (0.22, 0.41) | **<0.00001** | 0.29 (0.20, 0.38) | **<0.00001** |
| Average RNase P gene value | 0.40 (0.35, 0.45) | **<0.00001** | 0.39 (0.34, 0.44) | **<0.00001** |
| **Vaccinated individuals (complete primary series or booster dose at time of swab), adjusted for age, symptoms status, vaccination status, days since last COVID-19 vaccine dose, and average RNase P gene value**<br>**N=1,025 (Delta = 84, Omicron = 941)** |  |  |  |  |
| Variant (Omicron vs. Delta) | 1.26 (0.47, 2.05) | **0.001** | 0.87 (0.08, 1.67) | **0.03** |
| Age (years) | -0.003 (-0.03, 0.02) | 0.74 | -0.01 (-0.03, 0.01) | 0.48 |
| Symptoms (symptomatic vs. asymptomatic) | -1.27 (-2.86, -0.16) | **0.00001** | -1.41 (-1.93, -0.88) | **<0.00001** |
| Booster vaccination vs. Complete primary series vaccination | 0.71 (0.19, 1.22) | **0.007** | 0.50 (-0.55, 1.54) | **0.35** |
| Days since last COVID-19 vaccine dose | -0.002 (-0.005, -0.0004) | **0.02** | -0.001 (-0.006, 0.003) | 0.8287 |
| Average RNase P gene value | 0.37 (0.31, 0.43) | **<0.00001** | 0.38 (0.32, 0.44) | **<0.00001** |

\*Mean adjusted differences estimated using three multiple linear regression of average Orf1b Ct value on variant (Omicron vs. Delta) adjusted by covariates indicated in the table above. All regressions were restricted to Delta and Omicron cases detected using RHINOstic™ swabs (excludes 27 Delta and 42 Omicron cases detected using US Cotton #3 swabs).

### Intra-residence transmission

Among the 1,939 SARS-CoV-2 genomes, we identified 13 residences with multiple sequenced Delta cases and 136 residences with multiple sequenced Omicron cases. Phylogenetic and pairwise distance analyses of these genomes indicated that many cases within the same residence were likely the result of more than one introduction event. Thus, we restricted analysis to 78 clusters including 173 individuals with identical viral genomes within the same residence (N=25 residents for Delta, and 148 for Omicron).

Thirty individuals reported that symptoms began on the same day as another individual in the cluster and 53 collected their first positive sample on the same day as another individual in the cluster. All identical viral genomes within a single household were detected within a maximum serial interval of 15 days. Forty-four clusters included more than one symptomatic individual and more than one unique symptom onset date. Among these clusters, the median serial interval between symptom onset of the index and a subsequent case was longer for 8 subsequent cases in 7 Delta clusters (median 6 days, range [1-10]) compared to for 43 subsequent cases in 37 Omicron clusters (median 2 days, [1-9]) (P=0.021, **Supplemental Figure 1**).

### Genomic analysis

A phylogenetic tree shown in **Figure 3A** includes all 209 Delta genomes shown with 1,174 randomly selected genomes from samples collected in Washington state over the same time period. A phylogenetic tree containing all 1,939 sequenced viral genomes is shown in **Figure 3B**, illustrating rapid replacement of Delta by Omicron on the university campus in December 2021. Three monophyletic clusters containing exclusively or almost exclusively study genomes (N=35, 24, 66 total genomes and N=35, 23, 66 HCT genomes) are boxed in Figure 3A; approximately 60% of all study Delta genomes fall into one of these 3 groups. The maximum pairwise distance between two study Delta samples was 60, and average distance was 18.54. One hundred sixteen (56%) of these samples were genetically identical to at least one other study sample. The tree in **Figure 3C** includes all 1,730 Omicron genomes with 1,512 randomly selected genomes from samples collected in Washington state over the same time period. Relative to the Delta genomes, the study Omicron genomes are more evenly distributed throughout the tree, particularly genomes from samples collected in January and February. The maximum pairwise distance between two study Omicron samples was 89 and average distance was 7.10 (72 and 6.01, respectively, excluding BA.2 samples). One thousand thirty-nine (60%) of Omicron samples were genetically identical to at least one other study sample. Among the 1,730 sequenced Omicron samples, 24 were of the BA.2 lineage. The maximum pairwise distance among these was 9, and 19 (79%) were identical to at least one other study genome.

**Figure 3.**
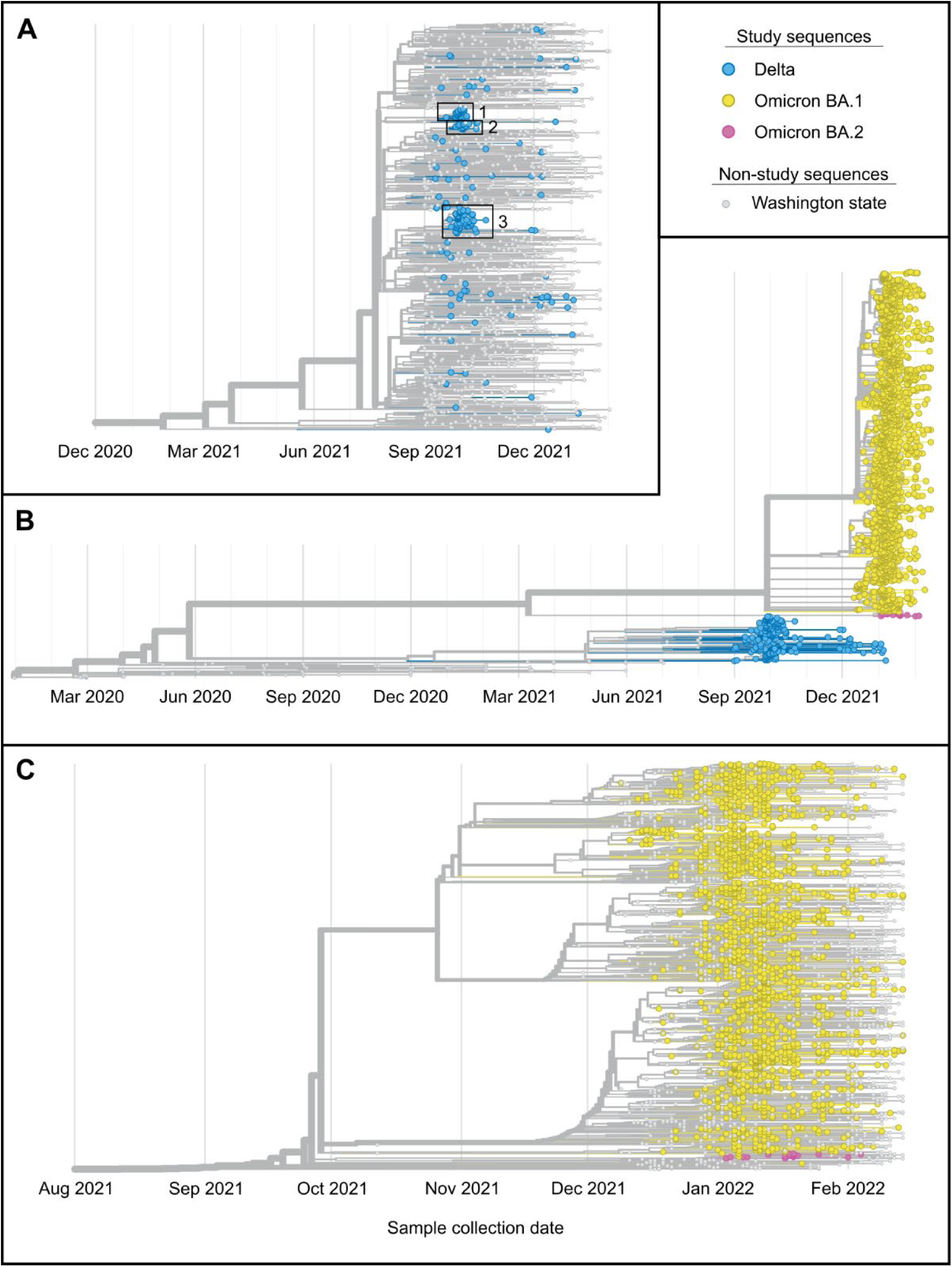
Phylogenetics of sequenced campus viral genomes. (A) Phylogenetic tree of 209 sequenced Delta samples collected on the UW campus and 1,174 randomly selected genomes from samples collected in Washington during the same time period. Three monophyletic clusters containing exclusively or almost exclusively study genomes are boxed and numbered. (B) Phylogenetic tree containing sequences for campus samples collected between September 4, 2021 and February 14, 2022 (N=1,939) plus the Wuhan/Hu-1 reference genome and approximately 100 GISAID Washington state genomes collected from March 2020 to August 2021. The tree also contains genomes for 94 samples collected in Washington state from March 2020 to August 2021 (gray nodes) and the Wuhan/Hu-1 reference genome (gray node, far left) for context. Delta variant campus genomes are in yellow, and Omicron variant genomes are in blue. (C) Phylogenetic tree of 1,730 sequenced Omicron samples collected on the UW campus plus the Wuhan-Hu-1 reference genome, and 1,512 randomly selected genomes for samples collected in Washington during the same time period.

To estimate the number of introduction events of Delta into the campus population needed to explain the sequenced samples, we created a phylogenetic tree including all sequenced Delta study samples and all publicly available genomes for Delta viruses collected in Washington state from September 1, 2021, to February 14, 2022, for a total of 209 genomes from our study and 15,406 Washington state genomes. By determining the likely classification of internal nodes as either campus or community-based, we estimated that the sequenced Delta samples resulted from 83 different introductions of the variant with 2.5 sequenced cases per introduction. We performed the same analysis for the sequenced Omicron samples using 14,359 publicly available Omicron genomes from samples collected in Washington state up to February 14, 2022. We estimate that 1,021 introduction events were necessary to explain the 1,730 sequenced Omicron cases, with 1.7 sequenced cases per introduction. We also assessed the Omicron BA.2 subvariant viruses separately. We created a tree containing the 24 BA.2 viral genomes generated from samples collected on campus plus 126 BA.2 genomes from samples collected in Washington up until February 14, 2022. We estimated that the 24 sequenced study cases resulted from 8 different introductions with 3.0 sequenced cases per introduction. To assess the accuracy of the Delta and Omicron introduction number estimates, we repeated these analyses using smaller pools of Washington state (non-study) genomes. This assessment showed that the estimate of Delta introduction events would be unlikely to change even if more Washington state genomes were available, though it was unclear if this was the case for the Omicron estimate (**Supplemental Figure 2**).

### Transmission dynamics

To quantify the degree to which each variant impacted on-campus transmission rates, we estimated variant-specific transmission dynamics following previously established methods^35^. Here we find that the R_t_ associated with the September to October Delta outbreak peaked at 1.8 (95% credible intervals [CI] 1.3-2.4) and declined rapidly below 1, while the R_t_ associated with the December to January Omicron outbreak peaked at 2.4 (95% CI 1.9-2.8) and declined below 1 over a longer period (**Figure 4**). These differences in R_t_ are reflected in the relative magnitudes of the September to October Delta outbreak compared to the December to January Omicron outbreak (Figure 4).

**Figure 4.**
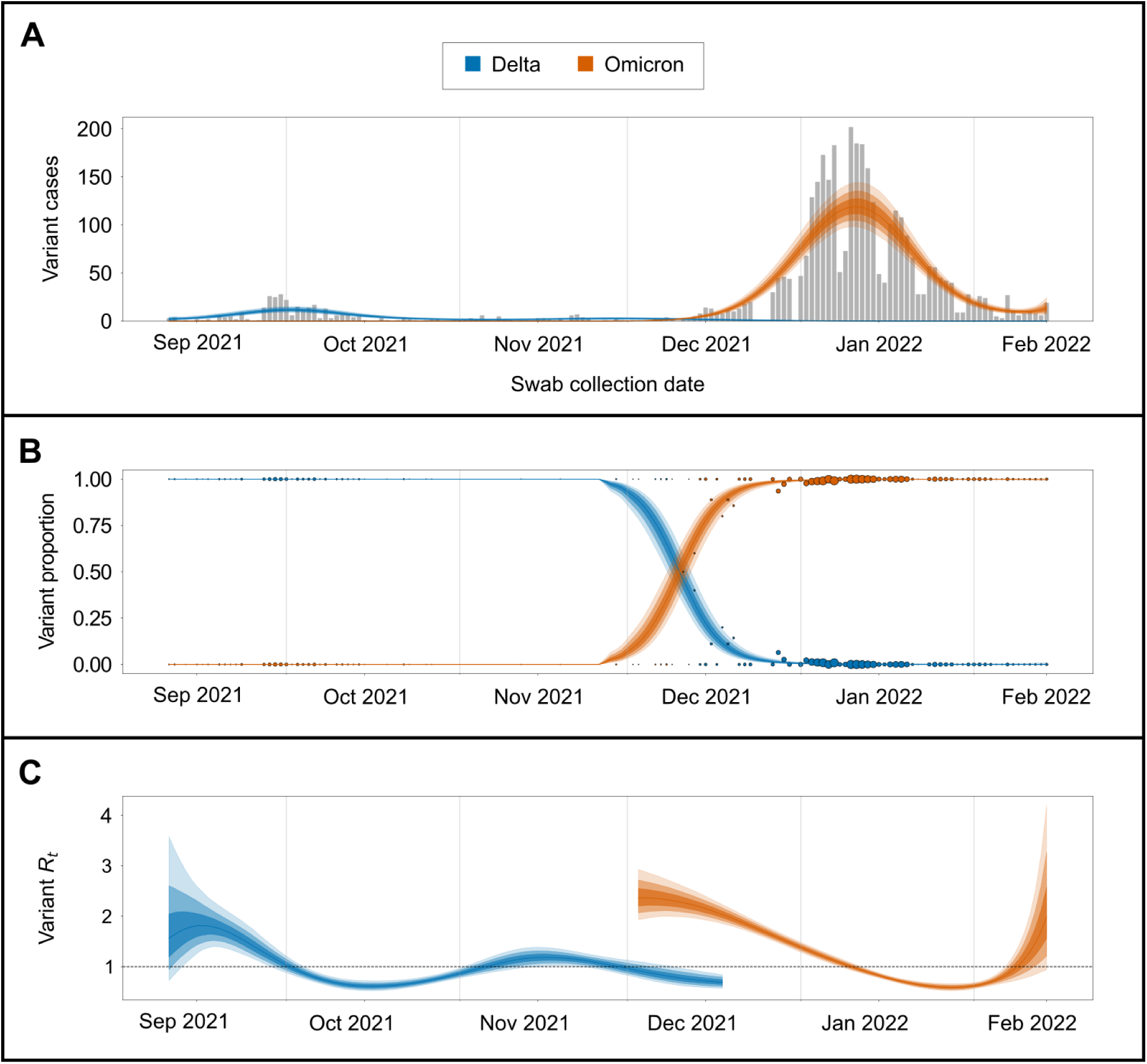
Transmission dynamics of Delta and Omicron over study period. (A) SARS-CoV-2 positive samples (gray bars) against posterior variant-specific incidence over the study period. (B) Observed variant proportion of sequenced positive SARS-CoV-2 samples against posterior variant proportion. Radius of points corresponds to count of sequenced samples for that day. (C) Posterior estimates of variant-specific effective reproduction numbers. Shaded intervals in all plots correspond to 50%, 80%, and 95% credible intervals.

## Discussion

In a large, urban university campus with widely available testing, stringent mitigation measures and near-universal vaccination, the Omicron variant rapidly displaced the Delta variant to become the predominant viral strain over a two-week period. Fever, myalgia, and chills were more commonly reported in Omicron cases and loss of taste and smell in Delta cases. Ct values were on average higher for Omicron cases. Using genomic analyses, we observed shorter serial intervals in case clusters and faster spread for Omicron relative to Delta. These findings highlight the importance of integrating genomic surveillance into university testing studies to better characterize VOC community spread.

Variants have continuously altered our understanding of SARS-CoV-2 genomic epidemiology. The adaptation of public health recommendations to this quickly changing landscape relies on rapid data collection, and university testing programs are uniquely positioned to collect data which may be more broadly representative of community dynamics than hospital-based surveillance strategies. Using symptom and exposure-based testing, we identified Omicron cases and characterized viral loads, serial intervals, and symptoms through daily online questionnaires in real-time as the first introductions of Omicron occurred. Prospective, longitudinal data collection from dormitories and other congregate settings offer an opportunity to understand transmission dynamics of viral infections within clusters. For example, traditional household studies including the Household Influenza Vaccine Evaluation^36^ and the Seattle Virus Watch^37^ have informed public health recommendations for influenza-related isolation and quarantine. Our university-based study with students residing in shared housing allowed for rapid data collection and decision-making around the evolving transmission dynamics of VOCs.

Our findings suggest a median serial interval of 2 days and 6 days among persons with Omicron and Delta, respectively. In contrast to other analyses examining serial intervals within households or other clusters^13–15,17,19^, we used viral genomic data to minimize confounding of the serial infection interval by co-incident exposures during periods of high community transmission. By using only identical genomes to calculate the intra-residence serial interval, we decreased the likelihood that clusters are the result of more than one index case (although we cannot eliminate this possibility). Our finding of reduced serial interval between index and subsequent household infections for Omicron compared to Delta cases is consistent with other studies in the US (median serial interval of 3 days for Omicron)^13^ and others in Europe and South Korea (reported mean serial intervals from 2.8-3.5 days for Omicron and 3-4.1 days for Delta)^16–19^. Our estimated median serial interval of 2 days for Omicron is lower than these studies and this may be due to our study population being on average younger and more highly vaccinated and only one other study using genomic sequencing to identify household transmission^16^.

Semiquantitative viral loads were lower for Omicron compared to Delta variant infections, supporting the theory that increased transmissibility of the Omicron variant is not due to viral load and in agreement with other studies in the setting of highly vaccinated populations and routine testing of asymptomatic and mildly ill individuals, including three other US universities and the US National Basketball Association’s occupational health program^9,12^. In contrast, a US study of hospital patients tested as part of routine clinical care and a Swiss study of symptomatic outpatients with only eighteen Omicron cases did not find a difference in viral load between Omicron and Delta variants^10,11^.

The Omicron variant swiftly replaced Delta on campus, despite high rates of vaccination and broad campus mitigation measures in place. Due to the availability of rapid whole genome sequencing^38,39^, we quickly identified the emergence of Omicron. The rapid rise of Omicron may have been facilitated by vaccine breakthrough cases and immune evasion associated with this variant, as reported in early Omicron studies^40–42^. Despite higher numbers of Omicron infection after vaccination, early household transmission reports show that individuals who received a booster dose had lower secondary attack rates, lower risk of transmission, and less secondary infections^14^. The pace of Omicron variant spread in this population, quantified as an increased R_t_ compared to Delta variant, exemplifies that SARS-CoV-2 outbreaks may continue to occur despite stringent public health interventions. To mitigate further waves of SARS-CoV-2 transmission, community-based genomic surveillance studies should be leveraged to guide policy and containment strategies. This reality, and a needed shift in the national COVID-19 strategy to focus on a “new normal” in which risk reduction and hospital capacity are prioritized, are essential as we transition to the next phases of the pandemic^43^.

Although SARS-CoV-2 variant classification may be achieved without full genome sequencing, generation of complete viral genomes provided additional insight into viral transmission on campus. Our genomic data suggest that within 2 months of the first detection of Omicron on campus, there were at least 1,000 distinct introductions of the variant, though our ability to precisely define the number of introduction events represented by the sequenced campus Omicron cases is limited likely due to the limited genomic variation among Omicron viruses and the fact that study genomes currently make up about 10% of the available Omicron genomes from Washington. This estimate does suggest that most Omicron introduction events resulted in a single sequenced case. Our analyses indicate that the same is true for Delta introduction events. However, for Delta, it was also clear that most sequenced cases were the result of introduction events that resulted in multiple cases, and that most on-campus SARS-CoV-2 cases due to Delta variant viruses were the result of campus-related transmission. It is particularly notable that most sequenced Delta cases were due to just one of three putative introduction events while the highest number of cases due to a single putative Omicron introduction event (for the analysis including all Washington state Omicron sequences) was 41 (or 2.4% of the total number of sequenced cases), which may suggest differences in patterns of Delta and Omicron transmission on campus. Unfortunately, the considerable degree of uncertainty in the Omicron phylogenetic tree limits our ability to directly compare transmission patterns of the two variants.

Our study limitations include the lack of routine surveillance testing of the entire campus population. Follow-up symptom data was missing for some individuals and therefore we do not know if some asymptomatic cases were pre-symptomatic. We rely on self-report of vaccine status and could not reference state registries. However, state registry data may be incomplete or delayed, especially for students from other states. A limitation of our Ct analysis is the change in swab type during the study which may impact viral load, and we therefore restricted our viral load analysis to only one swab type. We also did not account for repeat infections. Finally, this study included only people on a single university campus who participated in the research study, and who are on average, younger, healthier, and more educated than the general population.

In conclusion, we found rapid replacement of the SARS-CoV-2 Delta variant with the Omicron variant within a highly vaccinated university population. As we move into the next phases of the pandemic, real-time data around viral kinetics and genomic epidemiology of emerging variants will be important to guide our national strategies on mitigating respiratory virus spread.

## Data Availability

Participants of this study did not agree for their data to be shared publicly, and so supporting data is not available.

## Acknowledgements

We would like to thank the study participants. We also thank the UW Environmental Health & Safety COVID-19 Prevention & Response team including Katia Harb, Sheryl Schwartz, Natalie Thiel, and Kim Baker, UW administration and Incident Command leadership group (Margaret Shepherd, Josh Gana, Pamela Schreiber), Chu Lab, the Brotman Baty Institute, Husky Coronavirus Testing team, Dr. Timothy Uyeki, Dr. Anna Wald, and Dr. Roy Burstein. We acknowledge the authors, originating and submitting laboratories of the sequences from GISAID’s EpiCoV database, on which this research is based. An acknowledgments table is provided as Supplementary Data.

## SUPPLEMENTAL MATERIALS

**Supplemental Table 1.** Cycle threshold comparisons by Omicron lineage (BA.1 vs. BA.2).

|  | Mean unadjusted<br>difference in Orf1b Ct<br>value (95% CI) | p-value | Mean adjusted<br>difference in Orf1b Ct<br>value (95% CI)* | p-value |
| --- | --- | --- | --- | --- |
| **All Omicron positive individuals, adjusted for age, symptoms, and average RNase P gene value**<br>**N=1,688 (BA.1 = 1664, BA.2 = 24)** |  |  |  |  |
| Lineage (BA.1 vs. BA.2) | 0.47 (-0.94, 1.88) | 0.51 | -0.04 (-1.34, 1.26) | 0.95 |
| Age (years) | -0.02 (-0.04, 0.002) | 0.09 | -0.01 (-0.03, 0.003) | 0.11 |
| Symptoms (symptomatic vs.<br>asymptomatic) | -1.06 (-1.49, -0.63) | **<0.00001** | **-1.25 (-1.65, -0.85)** | **<0.00001** |
| Average RNase P gene<br>value | 0.38 (0.33, 0.42) | **<0.00001** | 0.38 (0.34, 0.43) | **<0.00001** |

\*Mean adjusted differences estimated using multiple linear regression of average Orf1b Ct value on lineage (BA.1 vs. BA.2) adjusted for age, symptoms, and average RNase P gene value. Regression was restricted to Omicron cases detected using RHINOstic™ swabs (42 Omicron cases detected using US Cotton #3 swabs).

**Supplemental Figure 1.**
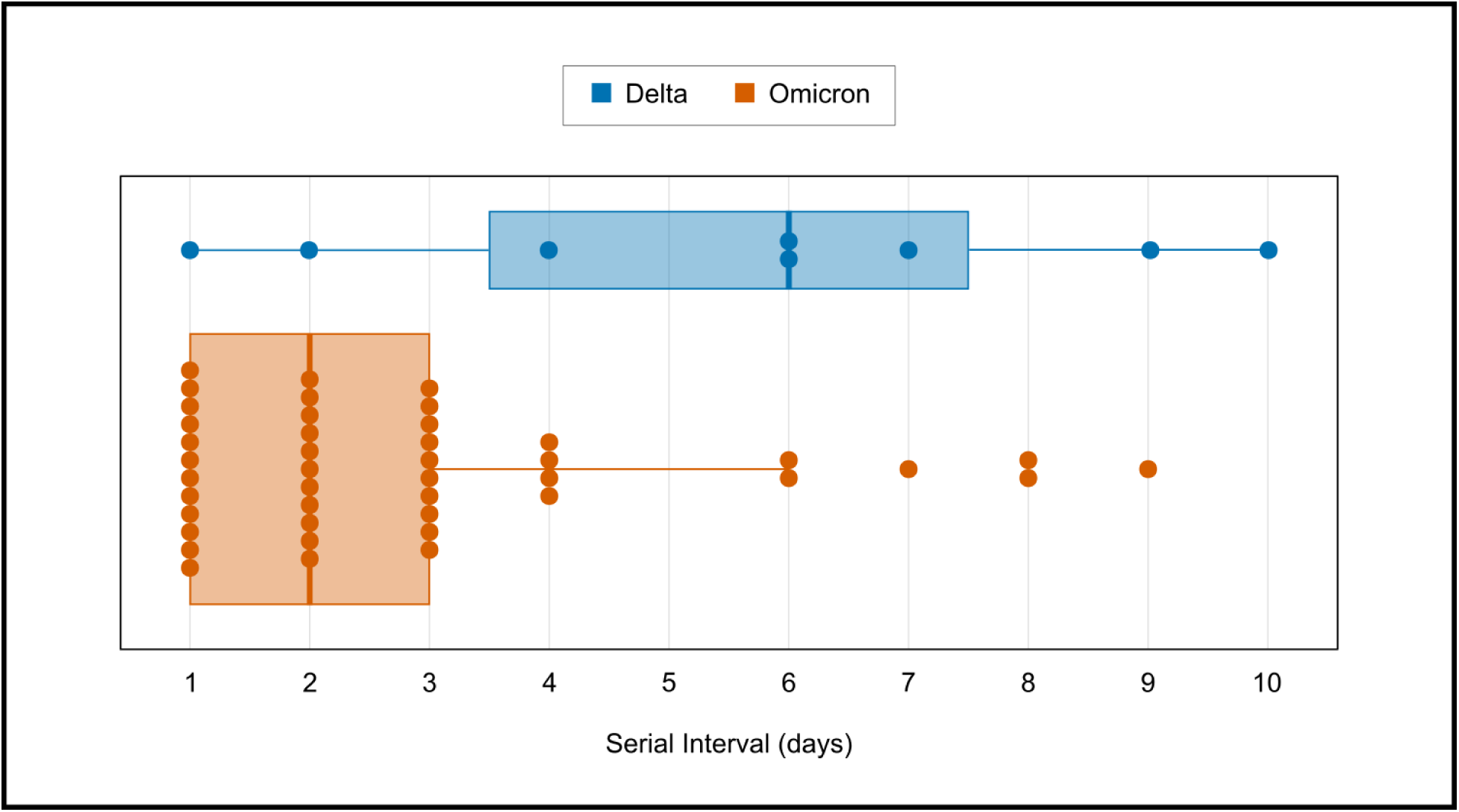
Serial intervals of Delta and Omicron transmission clusters. Serial intervals are displayed for each subsequent case in symptomatic clusters of SARS-CoV-2 positive individuals with identical genomes and sharing the same address. The primary case of each cluster was identified by the earliest symptom onset date within the cluster. Serial interval was defined as the number of days between symptom onset of a symptomatic primary case and each subsequent symptomatic case within the cluster.

**Supplemental Figure 2.**
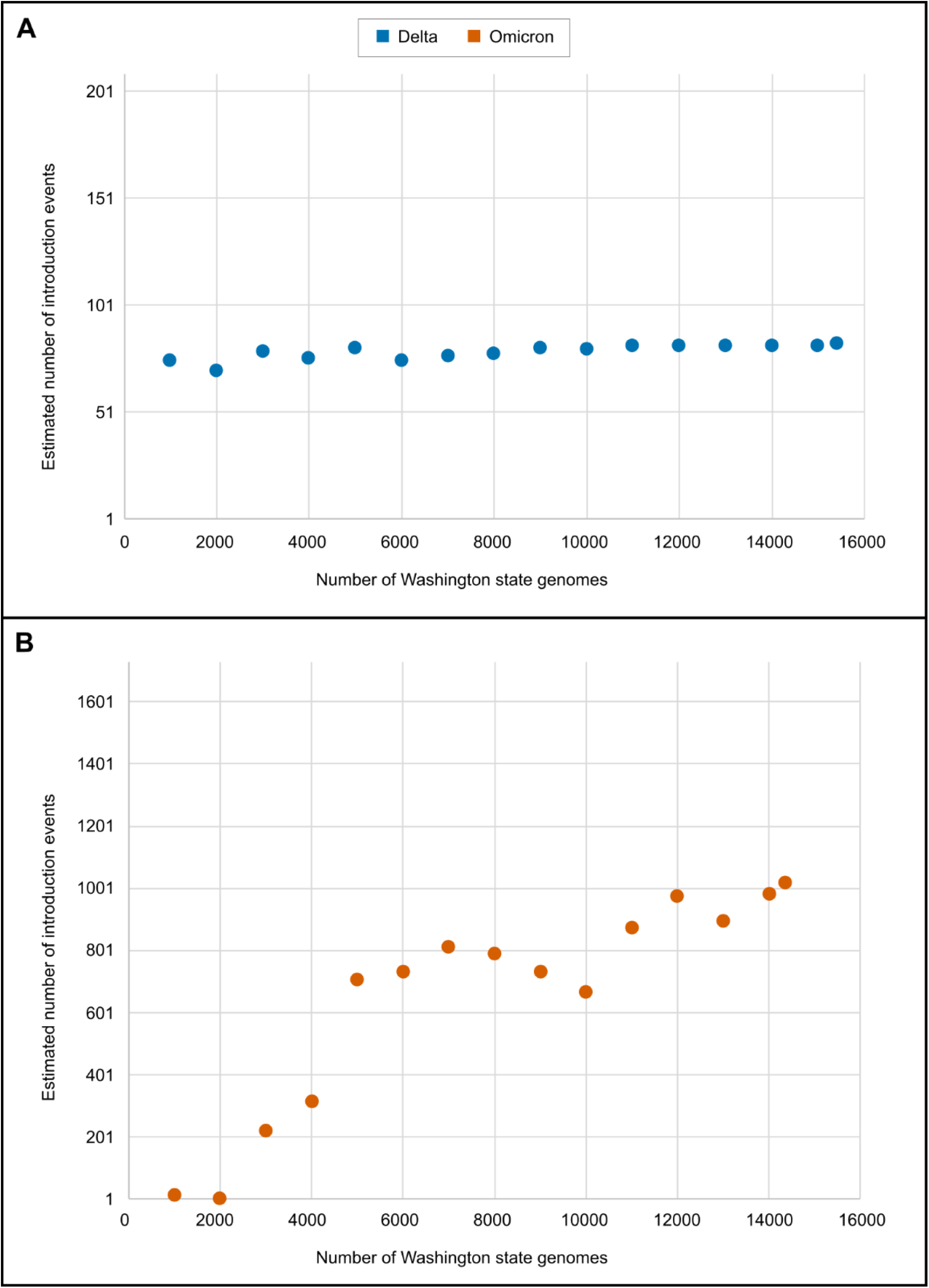
Accuracy analysis of number of Delta (A) and Omicron (B) introduction events onto campus represented by sequenced genomes. Sample size of the pool of non-study Washington state genomes used in each analysis is on the x-axis and the resulting estimate for the number of introduction event of each variant onto campus for each analysis is on the y-axis. In (A), the vertical axis varies from 1 to 209, the full range of possible values for the introduction number estimate for this variant. Similarly, in (B), the vertical axis varies from 1 to 1,730.

